# Fibrosis and liver inflammation are key regulators of α1-acid glycoprotein fucosylation

**DOI:** 10.1101/2023.11.14.23298443

**Authors:** Carlos Oltmanns, Birgit Bremer, Laura Kusche, Per Stål, Robin Zenlander, Jan Tauwaldt, Ingvar Rydén, Peter Påhlsson, Markus Cornberg, Heiner Wedemeyer

## Abstract

**Background and Aims:** There is an urgent need for new high-quality markers in the early detection of hepatocellular carcinoma (HCC). Åström et al. suggested that S2-bound α1-acid glycoprotein (AGP) might be a promising marker. Consequently, we evaluated S2-bound AGP for a predictive advantage in the early detection of HCC.

**Methods:** In a retrospective case-control study of patients chronically infected with hepatitis C virus (HCV) and treated with direct-acting antiviral agents (n=93), we measured S2-bound AGP using the HepaCheC® ELISA kit (Glycobond AB, Linköping, SE) at treatment start, end of treatment and follow-up (maximum: 78 months). Patients were retrospectively propensity score matched (1:2). 31 patients chronically infected with HCV developed HCC after sustained virological response while 62 did not. In addition, samples of HBV, MASLD and HCC from different etiologies patients were measured.

**Results:** S2-bound AGP elevation in HCC patients was confirmed. However, we did not observe a predictive advantage of S2-bound AGP in early detection of HCC during treatment and follow-up. Interestingly, S2-bound AGP levels correlated with aspartate aminotransferase (ρ=0.56, p=9.5×10^-15^) and liver elastography (ρ=0.67, p=2.2×10^-16^). Of note, S2-bound AGP decreased in patients chronically infected with HCV after treatment-induced clearance of HCV.

**Conclusion:** Fibrosis and liver inflammation are key regulators in the fucosylation of AGP. The potential role of S2-bound AGP as a novel tumor marker requires further investigation.

**Graphical abstract:** 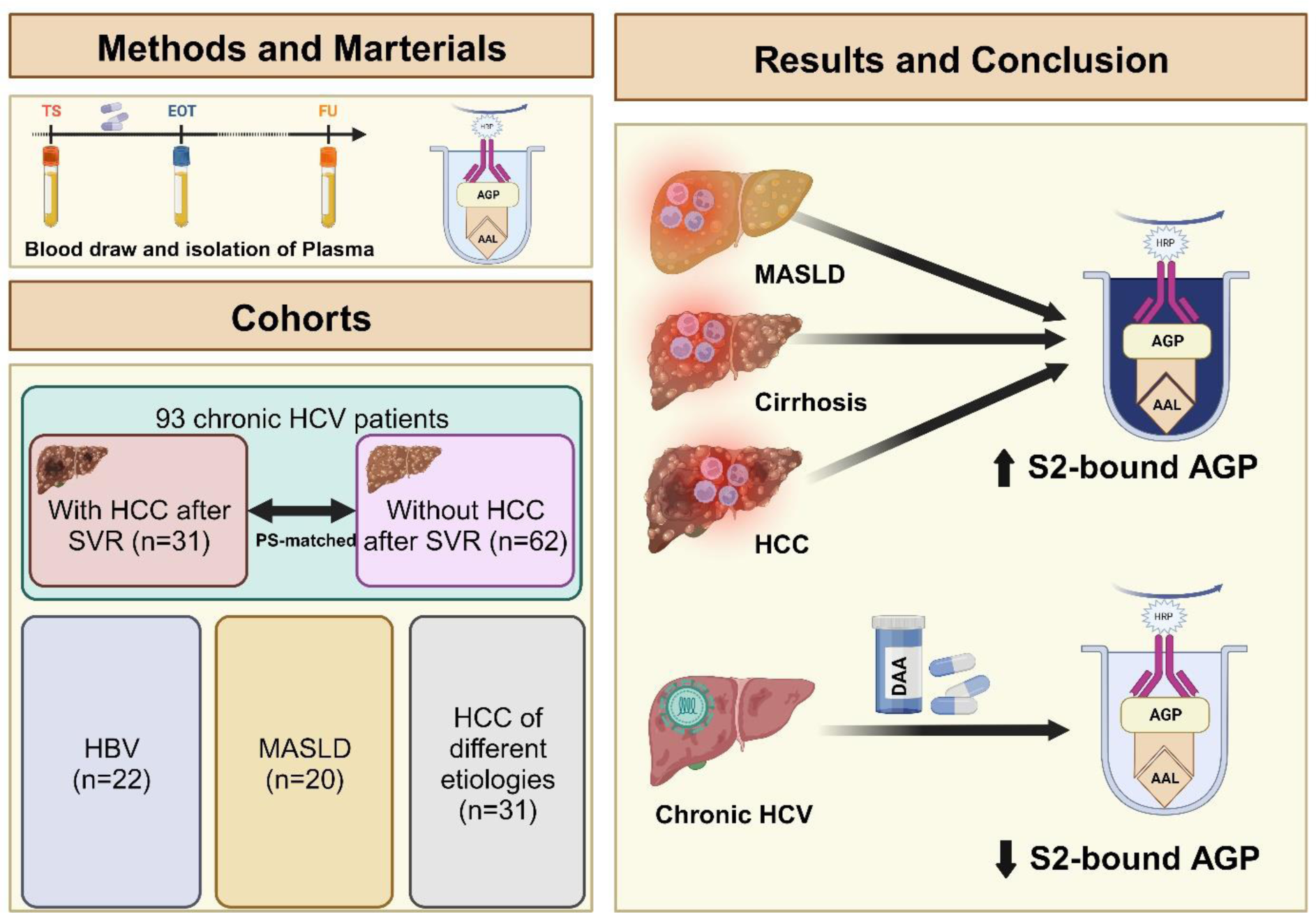

## Introduction

Hepatocellular carcinoma (HCC) is one of the leading causes of cancer-associated deaths worldwide [1]. We know that early detection of HCC can improve survival [2] [3]. Therefore, there is an urgent need to improve early detection [4]. As of today, ultrasound sonography with or without AFP is the gold standard for HCC surveillance [5]. Additional markers as AFP, AFP-L3 and DCP can improve sensitivity and specificity of HCC diagnosis. However, they do not allow an earlier diagnosis and their use has not been associated with an improved outcome [6] [7] [8].

Through the last decades, several groups showed that cancer development and severe liver disease affect serum glycoproteins [9]. Since AFP, AFP-L3 and DCP fall short of predicting HCC effectively, other glycoproteins have been studied extensively. Comunale et al. studied the fucosylation of alpha-1-antitrypsin (A1AT) and demonstrated that it is a potential biomarker candidate [10]. In contrast, Asazawa et al. studied fucosylated haptoglobin and showed a potentially higher sensitivity in detecting HCC than AFP alone [11]. Despite these efforts, none of these markers could meet the criteria for a high-quality predictive HCC marker and make it to the clinical stage.

Therefore, Åström et al. aimed to find a new HCC biomarker for early detection of HCC. They suggested that a reverse lectin based assay, measuring multifucosylated oligosaccharides, could differentiate between HCC and cirrhotic or non-cirrhotic hepatitis C virus (HCV) patients [12]. However, it remains unclear, whether S2-bound α1-acid glycoprotein (AGP) shows a predictive advantage and whether S2-bound AGP elevation is HCC specific or driven by inflammation and fibrosis.

We evaluated S2-bound AGP in a longitudinal retrospective case-control study consisting of patients, who underwent treatment of chronic hepatitis C virus infection with direct-acting antiviral agents (DAA). In addition, we demonstrate how liver inflammation and fibrosis affect S2-bound AGP levels in a real world setting. In the end, this study offers the opportunity to understand how inflammation and fibrosis might play a fundamental role in the fucosylation of glycoproteins.

## Materials and Methods

### Study population and design

We collected plasma samples from overall 93 patients chronically infected with hepatitis C virus. All patients were treated with DAAs since January 1, 2014. Thirty-one patients developed HCC after start of therapy with DAAs (HCV HCC cohort). In contrast, 62 matched controls did not develop liver cancer during the follow-up period (HCV Control cohort). All patients achieved sustained virological response (SVR) and the majority of patients suffered from cirrhosis. Patients that were lost to follow-up or did not achieve SVR (n=200), patients that did not have data available for the matching variables (n=26) and patients that were excluded during the matching process (n=480) were excluded from further analysis (Figure 1).

**Figure 1:**
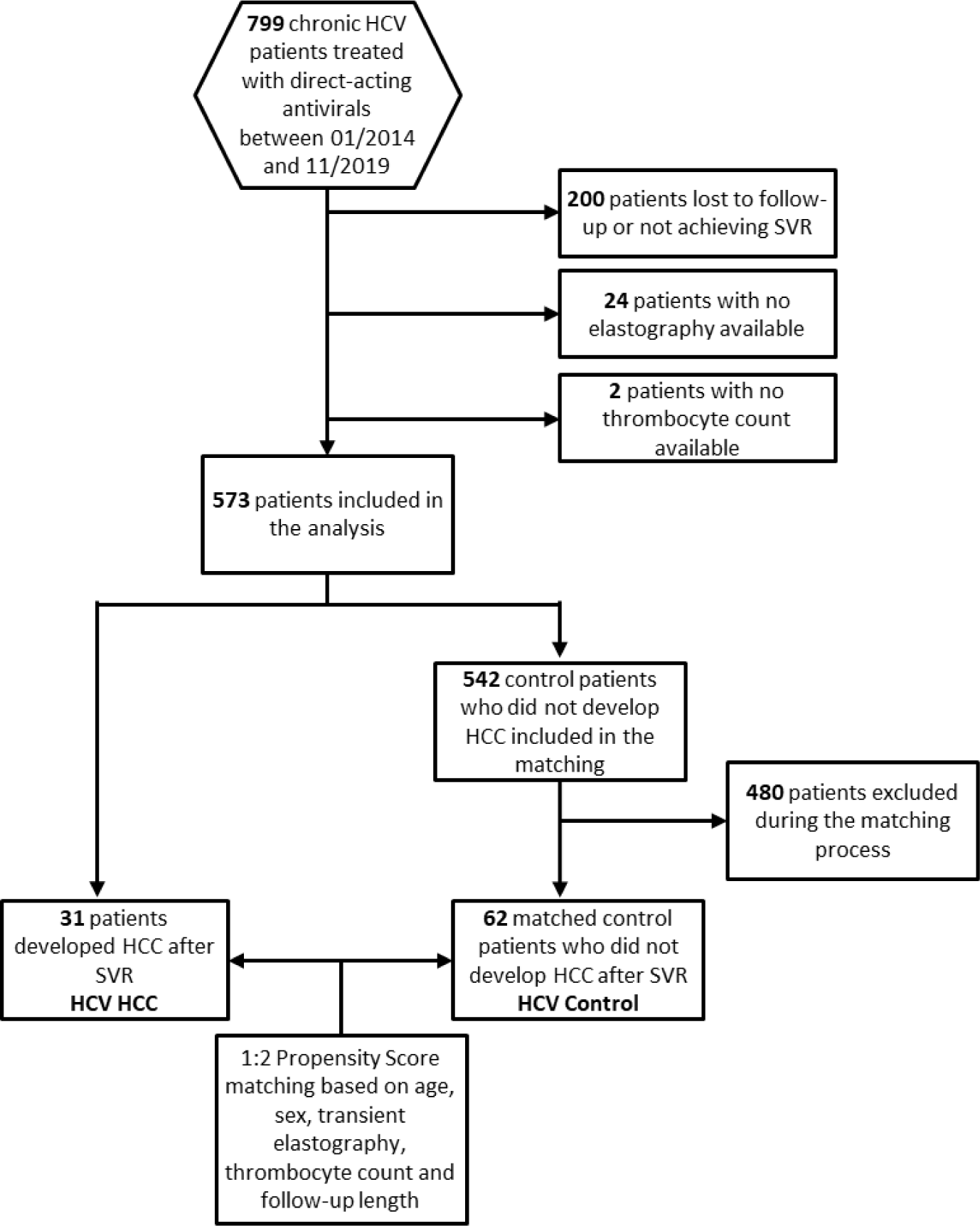
Inclusion and exclusion criteria for patients chronically infected with HCV.

We took samples at therapy start, end of treatment and at the time of HCC diagnosis (HCV HCC) or at the last available visit (HCV Control). All samples were collected in the outpatient clinic of Hannover Medical School between January 1, 2014 and March 31, 2021. We received written consent from all patients in advance.

In addition, we measured samples from different liver diseases independent of HCV infection. Three additional groups were measured, including patient chronically infected with hepatitis B virus (HBV) (n=22), patients with metabolic dysfunction-associated steatotic liver disease (MASLD) (n=20) and patients with HCC of different etiologies (n=31) (Suppl. table 1).

### Validation cohort

To validate our findings we analyzed a cohort of 173 patients from Sweden suffering from severe liver disease. One-hundred and two patients were diagnosed with liver cirrhosis whereas 71 patients were diagnosed with HCC. Etiologies of disease varied and most patients developed severe liver disease based on an HCV infection and/or high alcohol consumption. The HCC patients were either not treated for HCC (n=25), currently under treatment (n=23) or had completed the treatment (n=23).

### Cohort matching

We matched our 31 HCV HCC patients in a 1:2 propensity score (PS) matching with 62 HCV Control patients using R Statistical Software (Version 4.0.5.) and the MatchIt package [13]. In order to receive a robustly matched control cohort we chose nearest neighbor matching as the method of matching [14]. Our matching variables included sex, age, transient elastography, platelets and follow-up length.

### Sample preparation

Plasma was processed and then stored at a maximum temperature of -20° Celsius. We measured all samples using the HepaCheC® ELISA kit (Glycobond AB, Linköping, SE). The assay measures S2-bound AGP based on a reverse lectin ELISA [12]. We measured all samples according to the standard protocol of the manufacturer.

### Statistical analysis

We analyzed our data using R Statistical Software (Version 4.0.5.) and GraphPad Prism (Version 8.3.1.). Therefore, we created all graphs with the packages *ggplot2* and *ggpubr* or within GraphPad Prism. As statistical test, we performed “Mann-Whitney-U test” for comparison of independent sample groups. In addition, we calculated all correlations using Spearman’s rank correlation since the investigated correlations were non-linear. The graphical abstract was created using Biorender.com.

## Results

### Characteristics of the study cohort

Patient characteristics are shown in Table 1. In brief, patients chronically infected with HCV were middle-aged (HCV HCC: 59, HCV Control: 58.1) and mainly men (HCV HCC: 83.9% men, HCV Control 85.5% men). After PS matching, there were no differences between both groups regarding Child-Pugh score, HCV genotype, HCV-RNA, body mass index (BMI), platelets, aspartate aminotransferase (AST), alanine amino transferase (ALT), and albumin (Table 1A).

**Table 1:** Baseline characteristics of analyzed patients. Subgroups include patients who developed HCC after SVR (HCV HCC) and matched controls (HCV Control) (A). In addition, we analyzed patients with chronic HBV infection, MASLD and HCC (B). For continuous parameters, mean values and standard deviations and for categorical parameters, absolute and relative numbers are shown.

| **A** | **HCV HCC** | **HCV Control** | **P-value** | **Reference** |
| --- | --- | --- | --- | --- |
| **n** | 31 | 62 |  |  |
| **Age** | 59.0 (9.62) | 58.1 (9.56) |  |  |
| **Sex** |  |  |  |  |
| **m** | 26 (83.9%) | 53 (85.5%) |  |  |
| **f** | 5 (16.1%) | 9 (14.5%) |  |  |
| **HCV genotype** |  |  |  |  |
| **1** | 22 (71.0%) | 51 (82.2%) |  |  |
| **2** | 1 (3.2%) | 1 (1.6%) |  |  |
| **3** | 7 (22.6%) | 8 (12.9%) |  |  |
| **4** | 1 (3.2%) | 1 (1.6%) |  |  |
| **Child-Pugh Score** |  |  |  |  |
| **0** | 4 (12.9%) | 11 (17.7%) |  |  |
| **Child A** | 21 (67.7%) | 43 (69.4%) |  |  |
| **Child B** | 4 (12.9%) | 8 (12.9%) |  |  |
| **Child C** | 1 (3.2%) | 0 |  |  |
| **Elastography (kPa)** |  |  |  | 0-14.5 |
| **<14.5** | 5 (16.1%) | 10 (16.1%) |  |  |
| **14.5 - 25.0** | 3 (9.7%) | 13 (21.0%) |  |  |
| **>25.0** | 23 (74.2%) | 39 (62.9%) |  |  |
| **Body mass index** | 26.8 (2.69) | 27.9 (4.61) | 0.14 | 18.5-24.9 |
| **Platelets (Tsd/μL)** | 97.9 (49.3) | 96.1 (42.8) | 0.87 | 160-370 |
| **INR (Ratio)** | 1.27 (0.334) | 1.28 (0.348) | 0.86 | 0.9-1.25 |
| **AST (U/L)** | 97.5 (58.2) | 100 (55.3) | 0.83 | 0-35 (m) 0-31 (f) |
| **ALT (U/L)** | 92.7 (72.6) | 101 (65.3) | 0.61 | 0-45 (m) 0-34 (f) |
| **Albumin (g/L)** | 33.3 (6.12) | 35.5 (6.01) | 0.10 | 35-52 |
| **HCV-RNA (IU/mL)** | 1,240,000 (2,010,000) | 1,480,000 (1,760,000) | 0.58 | 0 |

| **B** | *HBV* | *MASLD* | *HCC* | *Reference* |
| --- | --- | --- | --- | --- |
| *n* | 22 | 20 | 31 |  |
| *Age* | 44.0 (11.6) | 67.6 (11.7) | 50.1 (11.4) |  |
| *Sex* |  |  |  |  |
| *m* | 11 (50.0%) | 7 (35.0%) | 25 (80.6%) |  |
| *f* | 11 (50.0%) | 13 (65.0%) | 6 (19.4%) |  |
| *Elastography (kPa)* | 4.7 (1.8) | 9.4 (3.3) | 35.8 (23.5) | 0-14.5 |
| *Platelets (Tsd/μL)* | 231 (52.5) | 237 (58.9) | 172 (99.4) | 160-370 |
| *INR (Ratio)* | 1.04 (0.36) | 1.01 (0.1) | 1.28 (0.65) | 0.9-1.25 |
| *AST (U/L)* | 26.2 (7.2) | 48.6 (18.0) | 79.3 (82.3) | 0-35 (m) 0-31 (f) |
| *ALT (U/L)* | 30.0 (12.9) | 87.0 (40.5) | 49.8 (47.3) | 0-45 (m) 0-34 (f) |

The HCV HCC group developed HCC after a median time of 28 months (Supplementary Figure 1), whereas the Control group did not develop HCC during follow-up (maximum follow-up: 78 months).

MASLD patients were overall older (MASLD: 67.6), whereas HBV and independent HCC patients showed younger age (HBV: 44.0, HCC: 50.1). Sex was equally distributed in the chronic HBV patients, MASLD patients were dominantly female (65% female), while HCC patients were mainly men (80.6%). AST and ALT were lower in the HCC patients than in the patients chronically infected with HCV but overall higher than in the HBV and MASLD patients. Elastography was similar in chronic HCV and HCC patients (Table 1B). Different etiologies of the independent HCC patients are presented in Supplementary Table 1.

### S2-bound AGP is elevated in HCC and chronic HCV patients

Confirming previous results, S2-bound AGP was increased in independent HCC patients with 16/31 measured samples above the proposed cut-off (=1). In contrast, S2-bound AGP was not elevated in chronic HBV (22/22 below cut-off) and MASLD (18/20 below cut-off) patients. We observed a highly significant difference between HCC patients and HBV or MASLD patients (HCC vs. HBV p<0.0001, HCC vs. MASLD p=0.0005). In addition, MASLD patients showed significantly higher S2-bound AGP levels than the HBV group (p=0.015).

Interestingly, patients chronically infected with HCV had highest S2-bound AGP levels, significantly higher than in the HBV (p<0.0001) and MASLD group (p<0.0001). We did not detect a significant difference in S2-bound AGP levels between HCC and chronic HCV patients. Overall, 49/93 patients chronically infected with HCV showed increased S2-bound AGP levels at baseline, which is similar to the HCC group (Figure 2).

**Figure 2:**
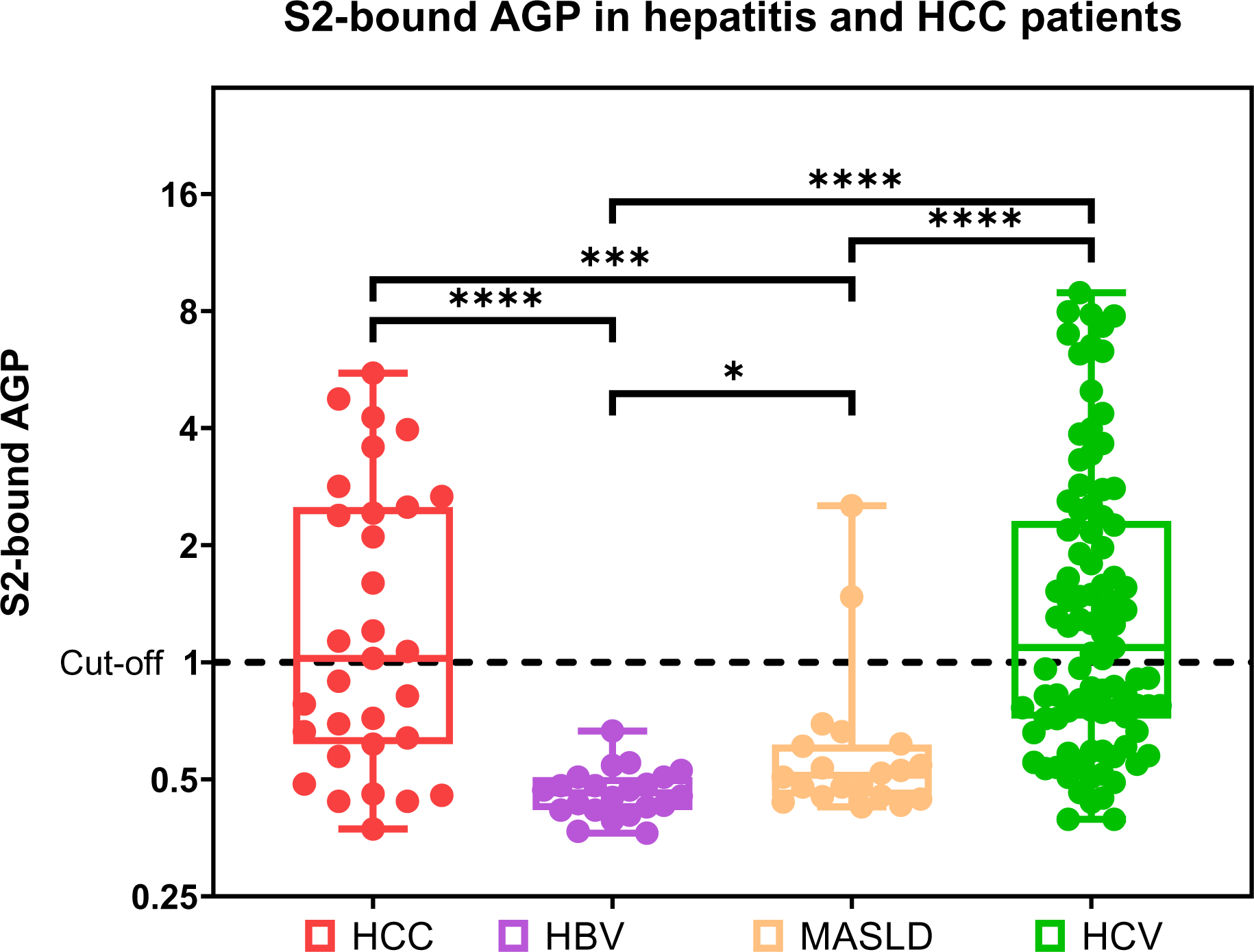
Concentration of S2-bound AGP in different groups of hepatitis and HCC patients. For patients chronically infected with HCV (green), baseline results are presented. Other groups included patients with chronic HBV infection (purple), patients with metabolic dysfunction-associated steatotic liver disease (orange) and patients with hepatocellular carcinoma (red). Shown are the median values for each group and the maximum and minimum values.

### S2-bound AGP correlates with AST and elastography

We next investigated to what extent markers of liver inflammation and fibrosis may correlate with S2-bound AGP. Indeed, both AST (ρ=0.56, p=9.5×10^-15^) and elastography (ρ=0.67, p=2.2×10^-16^) correlated with S2-bound AGP. Of note, a similar trend was found in subgroups including the patients chronically infected with HCV (AST: ρ=0.3, p=0.0047, elastography: ρ=0.36, p=0.00039) and HCC patients (AST: ρ=0.42, p=0.022) (Figure 3, Supplementary Figure 2).

**Figure 3:**
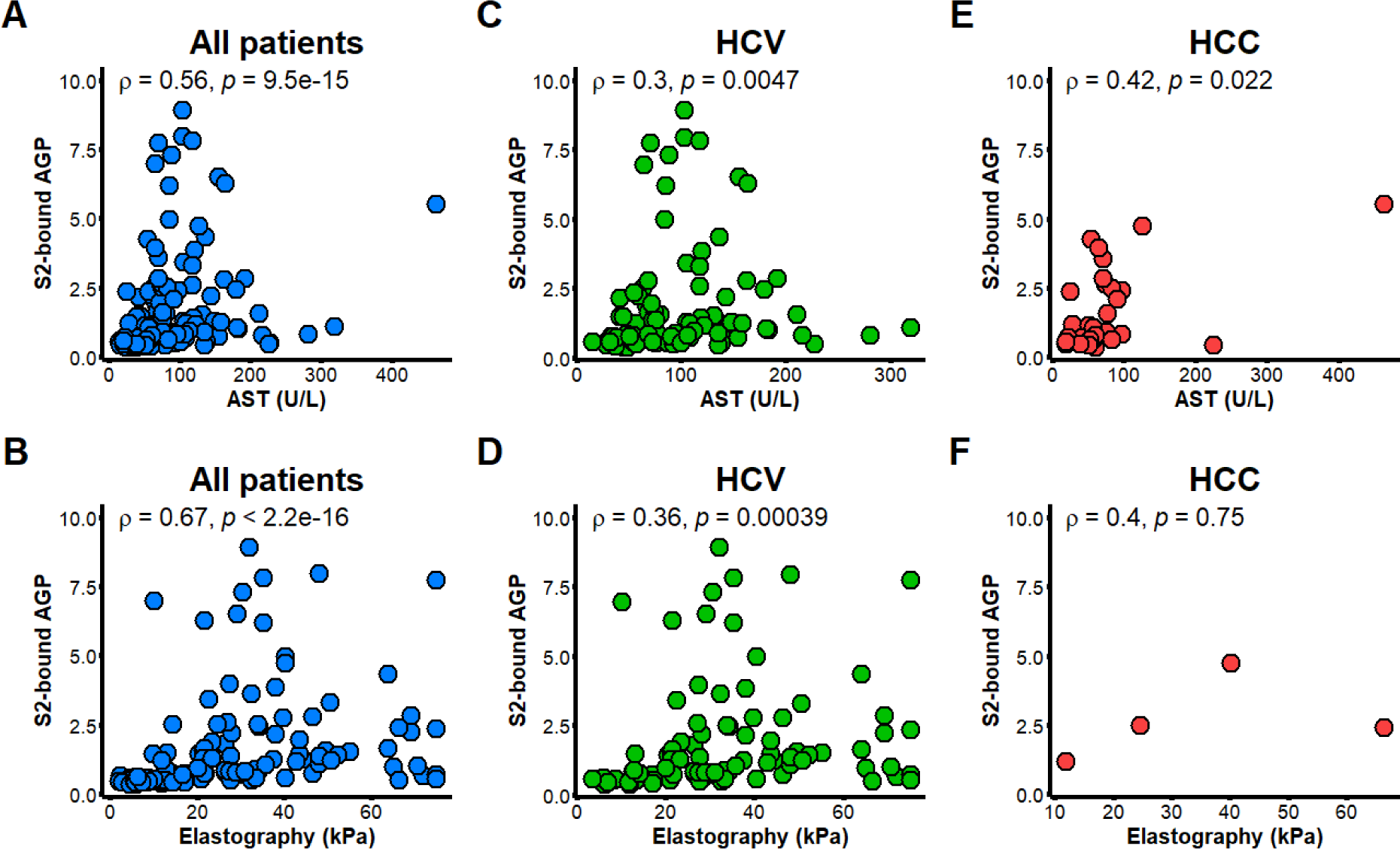
Concentration of S2-bound AGP correlated with clinical parameters (AST and elastography). Correlations are shown for all patients, including patients chronically infected with HCV at baseline (A-B), patients chronically infected with HCV at baseline (C-D), and HCC patients from different etiologies (E-F). Spearman’s Rank Correlation was used to calculate all correlations.

### S2-bound AGP correlates with AST and ALT in the validation cohort

To validate our findings we analyzed whether we could observe similar trends in the cohort of Swedish patients suffering from liver disease. Also here, we detected a correlation of S2-bound AGP with AST (ρ=0.35, p=3.1×10^-6^). In addition, there was also a weak correlation with ALT (ρ=0.21, p=6.2×10^-3^) (Figure 4).

**Figure 4:**
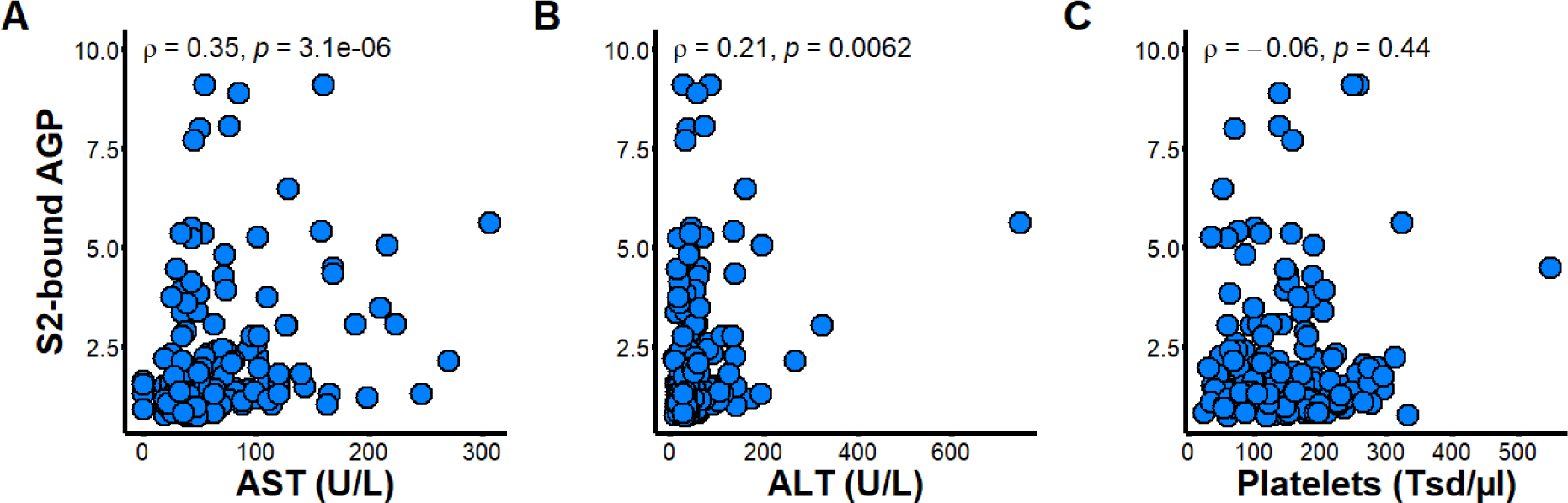
Concentration of S2-bound AGP correlated with clinical parameters (AST, ALT and Platelets) in a Swedish cohort of patients with severe liver disease. Spearman’s Rank Correlation was used to calculate all correlations.

### S2-bound AGP decreases in patients chronically infected with HCV after treatment

Building on top of these results, we analyzed the patients chronically infected with HCV longitudinally, measuring S2-bound AGP levels at baseline, at end of antiviral treatment and at long-term follow-up. The HCC and Control group both showed increased S2-bound AGP levels at baseline. Furthermore, S2-bound AGP levels decreased over time after successful treatment of the chronic HCV infection. We did not detect any significant difference between the patients that developed HCC after SVR and the patients from the Control group that did not develop HCC (HCC detection: p=0.10). This hold true during the complete course of treatment and follow-up (Figure 5).

**Figure 5:**
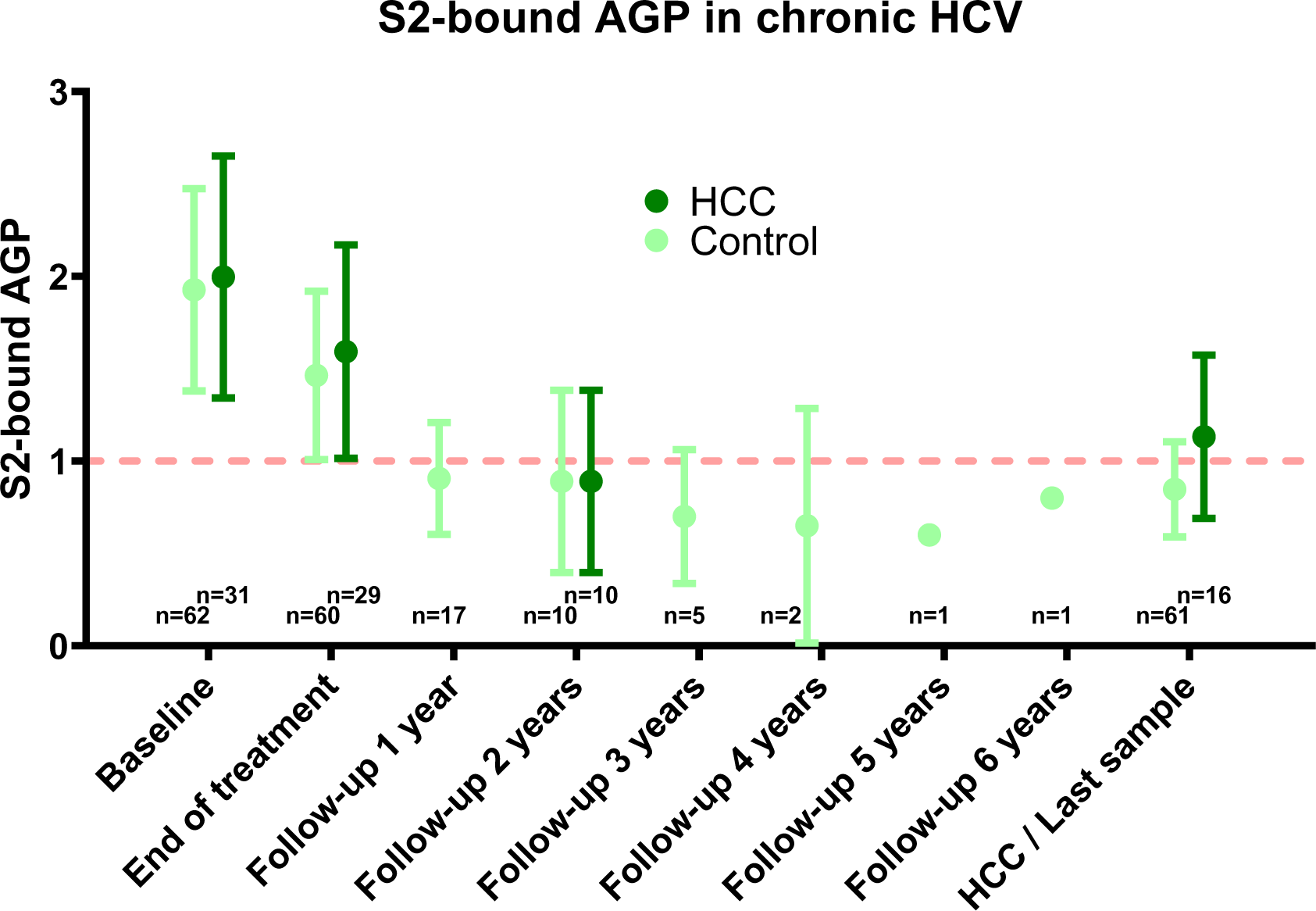
Longitudinal analysis of S2-bound AGP in patients chronically infected with HCV. Patients that developed HCC after SVR (dark green) are compared with the PS-matched control group (light green). Shown are the median values for each time point and the 95% confidence interval.

## Discussion

Despite decades of research effort, there is still a desperate need for predictive tumor markers to diagnose HCC early and with high precision [5]. In this study, we followed the hypothesis, that S2-bound AGP could be useful as a novel biomarker for early detection of HCC. Indeed, we confirmed that S2-bound AGP levels are elevated in half of the patients diagnosed with HCC, which is overall higher than traditional biomarkers, including AFP. However and surprisingly, the frequency of S2-bound elevation was similar in patients chronically infected with HCV. This suggests that non-specific inflammation and fibrosis may drive S2-bound AGP elevation. Therefore, S2-bound AGP is not a suitable candidate to predict development of hepatocellular carcinoma, specifically in patients that achieved SVR after chronic HCV infection.

The concentration of S2-bound AGP was associated with AST levels in two independent cohorts of patients. Although the association was evident in HCV, it was not statistically significant in the HBV or MASLD subgroups, where the levels of AST were lower than in the HCV group. However, we do not believe that the association of S2-bound AGP and AST is a HCV specific effect, nor do we believe that HCV infection or replication are specific drivers of AGP fucosylation. Interestingly, there was a significant reduction of S2-bound AGP levels after patients chronically infected with HCV successfully cleared the virus. This is in line with previous reports of traditional tumor markers that are also decreasing after SVR [15] [16]. However, this decline was much more prominent in our data on S2-bound AGP, suggesting a direct effect of chronic liver inflammation on fucosylation of AGP. Subsequently, most patients showed normalized liver enzymes and decreased levels of S2-bound AGP at follow-up compared to baseline.

In addition, the entity of liver disease may also play an important role in the assessment of new biomarkers. Whereas in patients chronically infected with HCV we observed high levels of S2-bound AGP, there were significantly lower levels in patients that were chronically infected with HBV or suffered from MASLD. At the same time, we observed lower levels of inflammation indicated by AST and ALT in these patients. However, our results are in line with previous findings on another tool for early detection of HCC, the GALAD score. Best et al. provided evidence that the entity of liver disease influences specificity and sensitivity of the GALAD score [17].

In the longitudinal cohort of patients chronically infected with HCV, S2-bound AGP did not show a diagnostic advantage in early detection of the HCC. Based on a previous study, Åström et al. suggested that S2-bound AGP could be specific for HCC [12]. While we see S2-bound AGP elevation in HCC patients in general, we could not see a significant difference between patients chronically infected with HCV and recently detected HCC and the HCV control group. Liver enzymes were elevated in the HCC patients of different entities too, leading us to the conclusion that general inflammation and fibrosis of the liver may influence previous study results.

Our results underline the challenges we are still facing in development of a high-quality predictive tumor marker in the HCC field [18]. All previous studies have failed to produce markers that can detect HCC reliably [19] [20]. Recently, Tayob et al. showed in an American Phase 3 biomarker study that GALAD indeed increases sensitivity for early detection of HCC but goes along with a significant false-positive rate of around 25 percent [8]. Therefore, it may be worth considering new approaches, such as using patients’ biological age as a predictive tool [21]. Finally, screening and evaluation of more promising candidates but also combination of different approaches might improve results for biomarker studies in the future.

Limits of our study include the relatively small number of patients in a retrospective study design from different cohorts. On the contrary, we were able to choose a well-matched cohort, allowing us to compare patients chronically infected with HCV that developed HCC to those that did not develop cancer. The close follow-up of patients after SVR allowed us to analyze pre-HCC-diagnosis-samples, bolstering our ability to evaluate, whether there is a predictive advantage of S2-bound AGP.

S2-bound AGP is affected by liver inflammation and does not predict HCC reliably on its own. Since the challenge of early HCC detection seems to be more complex, future research efforts should focus even more on a combination of markers [18]. Pattern recognition in multi-omics approaches and high throughput technologies in patients that develop HCC after chronic viral infection might play an important role. Fibrosis and inflammation of the liver influence S2-bound AGP and may influence fucosylation in other markers as well. Therefore, future studies on fucosylation should consider both to discriminate better between HCC-specific and non-specific signals.

## Compliance with Ethical Standards

### Informed consent and approval by the local ethics committee

This article does not contain any studies with animals performed by any of the authors. The study protocol conformed to the ethical guidelines of the Declaration of Helsinki and the local ethics committee approved this study a priori (No. 9474_BO_K_2020). Informed consent was obtained from all individual participants included in the study.

## Author contributions

HW conceived the project and coordinated the analyses. CO, BB and HW designed the experiments. CO, LK, JT, PP, IR, MC and HW drafted the manuscript. MC, PS, RZ and HW were involved in recruitment of patients. JT was involved in creation of the clinical cohort and BB helped acquiring the data. CO acquired and analyzed the data supervised by HW. All authors read and approved the manuscript.

## Abbreviations

A1AT: Alpha-1 antitrypsin
AFP: Alpha-fetoprotein
AFP-L3: Lectin-reactive alpha-fetoprotein
AGP: α1-acid glycoprotein
ALT: Alanine amino transferase
AST: Aspartate aminotransferase
DAA: Direct-acting antiviral agents
DCP: Des-gamma-carboxy prothrombin
GALAD: Gender, age, AFP-L3, AFP and DCP
HBV: Hepatitis B virus
HCC: Hepatocellular carcinoma
HCV: Hepatitis C virus
MASLD: Metabolic-dysfunction associated steatotic liver disease
PS: Propensity score
SVR: Sustained virological response

## Data Availability

All data produced in the present study are available upon reasonable request to the authors

## Supplementary material

**Supplementary Table 1:** Etiology of HCC in HCC group.

| Cause of HCC | Number of patients | Percentage |
| --- | --- | --- |
| HBV | 2 | 6.5% |
| HCV | 6 | 19.4% |
| HDV | 3 | 9.7% |
| MASLD | 4 | 12.9% |
| Cryptogenic fibrosis / cirrhosis | 2 | 6.4% |
| Hemochromatosis | 1 | 3.2% |
| Ethyl toxic | 6 | 19.4% |
| Ethyl toxic / viral hepatitis | 3 | 9.7% |
| Ethyl toxic / MASLD | 1 | 3.2% |
| Not available | 3 | 9.7% |

**Supplementary Figure 1:**
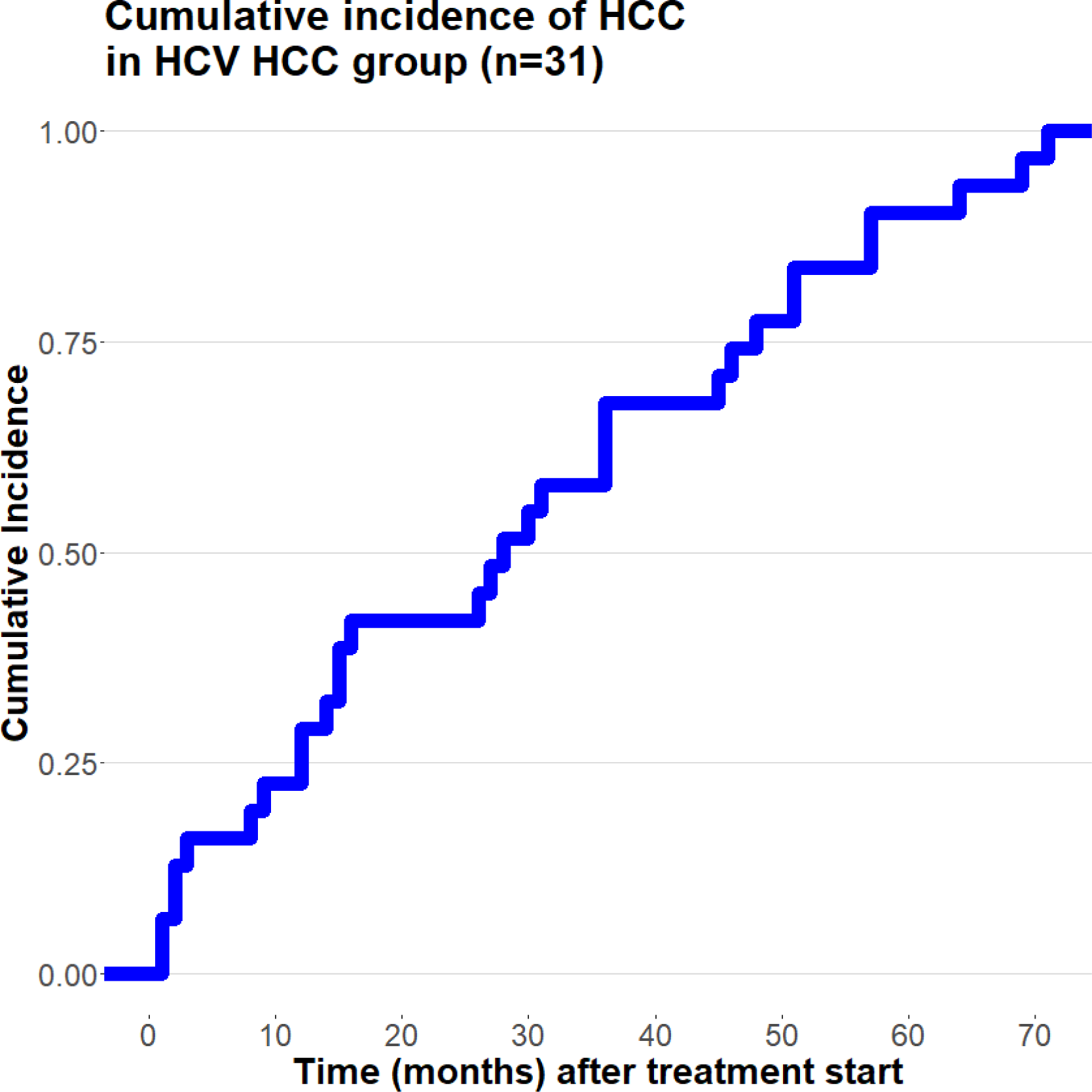
Cumulative incidence of HCC in HCV HCC group after treatment initiation.

**Supplementary Figure 2:**
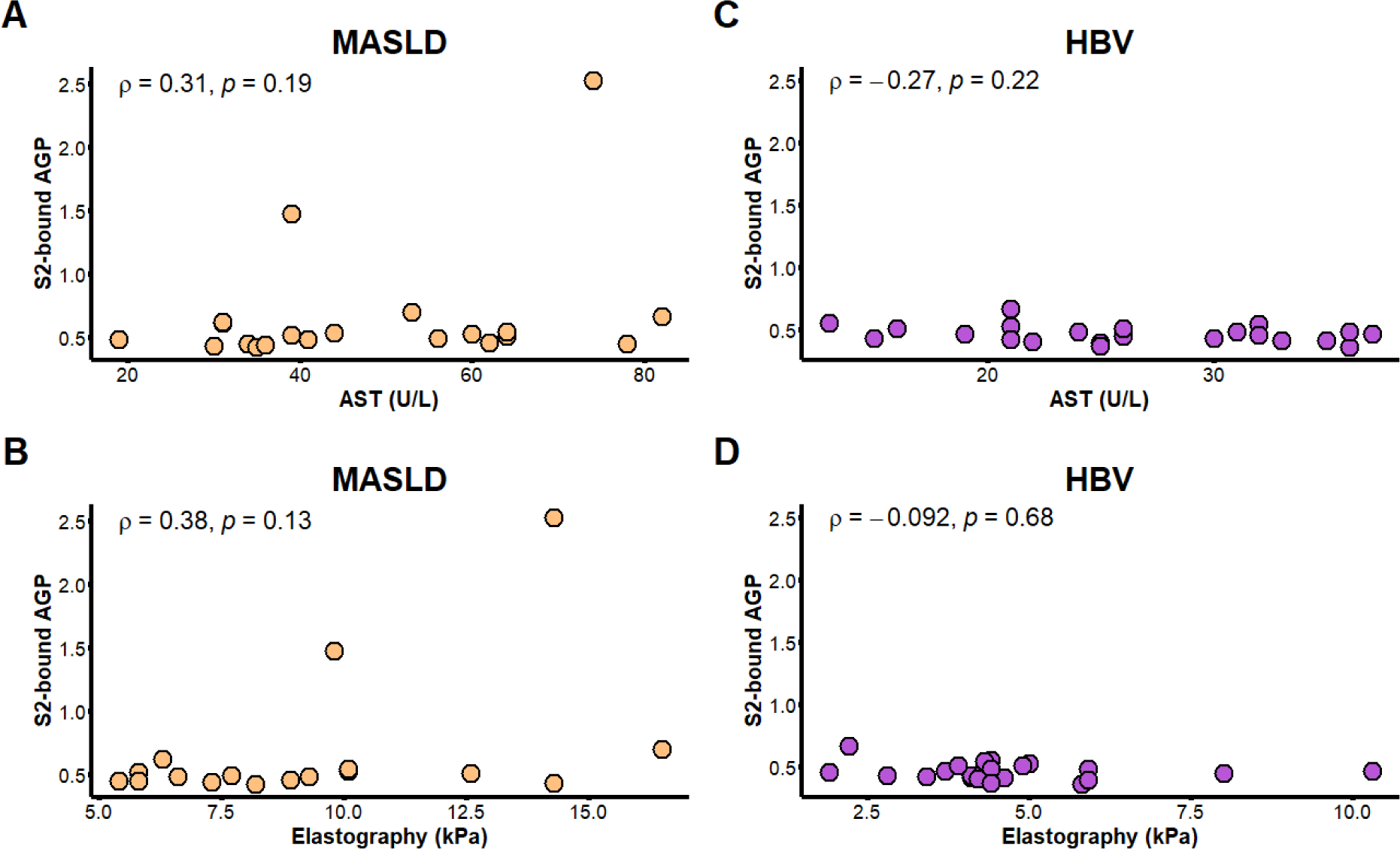
Concentration of S2-bound AGP correlated with clinical parameters (AST and elastography). Correlations are shown for MASLD (A-B) and chronic HBV (C-D) patients. Spearman’s Rank Correlation was used to calculate all correlations.

## References

[1] Sung H, Ferlay J, Siegel RL, Laversanne M, Soerjomataram I, Jemal A, et al. Global Cancer Statistics 2020: GLOBOCAN Estimates of Incidence and Mortality Worldwide for 36 Cancers in 185 Countries. CA: a cancer journal for clinicians 2021; 71:209–249.

[2] Singal A G, Zhang E, Narasimman M, Rich NE, Waljee AK, Hoshida Y et al. HCC surveillance improves early detection, curative treatment receipt, and survival in patients with cirrhosis: A meta-analysis. Journal of hepatology 2022; 77:128–139.

[3] Singal A G, Pillai A, Tiro J. Early Detection, Curative Treatment, and Survival Rates for Hepatocellular Carcinoma Surveillance in Patients with Cirrhosis: A Meta-analysis. PLoS Medicine 2014; 11:e1001624.

[4] Nault J, Villanueva A. Biomarkers for Hepatobiliary Cancers. Hepatology 2020; 73:115.

[5] Singal A G, Lampertico P, Nahon P. Epidemiology and surveillance for hepatocellular carcinoma: New trends. Journal of hepatology 2020; 72:250–261.

[6] Lersritwimanmaen P, Nimanong S. Hepatocellular Carcinoma Surveillance: Benefit of Serum Alfa-fetoprotein in Real-world Practice. Euroasian Journal of Hepato-Gastroenterology 2018; 8:83–87.

[7] Tzartzeva K, Obi J, Rich NE, Parikh ND, Marrero JA, Yopp A et al. Surveillance Imaging and Alpha Fetoprotein for Early Detection of Hepatocellular Carcinoma in Patients With Cirrhosis: A Meta-analysis. Gastroenterology (New York, N.Y. 1943) 2018; 154:1706–1718.e1.

[8] Tayob N, Kanwal F, Alsarraj A, Hernaez R, El-Serag HB. The Performance of AFP, AFP-3, DCP as Biomarkers for Detection of Hepatocellular Carcinoma (HCC): A Phase 3 Biomarker Study in the United States. Clinical Gastroenterology and Hepatology 2023; 21:415–423.e4.

[9] Naitoh A, Aoyagi Y, Asakura H. Highly enhanced fucosylation of serum glycoproteins in patients with hepatocellular carcinoma. Journal of gastroenterology and hepatology 1999; 14:436–445.

[10] Comunale M A, Rodemich-Betesh L, Hafner J, Wang M, Norton P, Di Bisceglie AM et al. Linkage Specific Fucosylation of Alpha-1-Antitrypsin in Liver Cirrhosis and Cancer Patients: Implications for a Biomarker of Hepatocellular Carcinoma. PLoS ONE 2010; 5:e12419.

[11] Asazawa H, Kamada Y, Takeda Y, Takamatsu S, Shinzaki S, Kim Y et al. Serum fucosylated haptoglobin in chronic liver diseases as a potential biomarker of hepatocellular carcinoma development. Clinical Chemistry and Laboratory Medicine (CCLM) 2015; 53:95–102.

[12] Åström E, Stål P, Zenlander R, Edenvik P, Alexandersson C, Haglund M et al. Reverse lectin ELISA for detecting fucosylated forms of α1-acid glycoprotein associated with hepatocellular carcinoma. PLoS ONE 2017; 12:e0173897.

[13] Ho D E, Imai K, King G, Stuart EA. MatchIt: Nonparametric Preprocessing for Parametric Causal Inference. Journal of statistical software 2011; 42.

[14] Randolph J J, Austin KF, Manuel K, Balloun JL, Randolph JJ;, Falbe K; et al. A Step-by-Step Guide to Propensity Score Matching in R. Practical Assessment, Research, and Evaluation 2014; 19.

[15] Fouad R, Elsharkawy A, Abdel Alem S, El Kassas M, Alboraie M, Sweedy A et al. Clinical impact of serum α-fetoprotein and its relation on changes in liver fibrosis in hepatitis C virus patients receiving direct-acting antivirals. European journal of gastroenterology & hepatology 2019; 31:1129–1134.

[16] Nguyen K, Jimenez M, Moghadam N, Wu C, Farid A, Grotts J et al. Decrease of Alpha-fetoprotein in Patients with Cirrhosis Treated with Direct-acting Antivirals. Journal of Clinical and Translational Hepatology 2017; XX:1.

[17] Schotten C, Ostertag B, Sowa J, Manka P, Bechmann LP, Hilgard G et al. GALAD Score Detects Early-Stage Hepatocellular Carcinoma in a European Cohort of Chronic Hepatitis B and C Patients. Pharmaceuticals (Basel, Switzerland) 2021; 14:735.

[18] Wang W, Wei C. Advances in the early diagnosis of hepatocellular carcinoma. Genes & Diseases 2020; 7:308–319.

[19] Nault J, Villanueva A. Biomarkers for Hepatobiliary Cancers. Hepatology 2020; 73:115.

[20] Piñero F, Dirchwolf M, Pessôa MG. Biomarkers in Hepatocellular Carcinoma: Diagnosis, Prognosis and Treatment Response Assessment. Cells 2020; 9.

[21] Oltmanns C, Liu Z, Mischke J, Tauwaldt J, Mekonnen YA, Urbanek-Quaing M et al. Reverse inflammaging: Long-term effects of HCV cure on biological age. J Hepatol 2023; 78:90–98.

